# An observational study on imported COVID-19 cases in Hong Kong during mandatory on-arrival hotel quarantine

**DOI:** 10.1101/2022.08.09.22278572

**Authors:** Mario Martín-Sánchez, Peng Wu, Dillon C. Adam, Bingyi Yang, Wey Wen Lim, Yun Lin, Eric H. Y. Lau, Sheena G. Sullivan, Gabriel M. Leung, Benjamin J. Cowling

**Author notes:** **Corresponding author:** Peng Wu.

## Abstract

**Background:** Hong Kong has enforced stringent travel restrictions particularly for inbound travellers since the emergence of SARS-CoV-2. Understanding the characteristics of imported COVID-19 cases is important for establishing evidence-based control measures.

**Methods:** We conducted a retrospective cohort study to summarise the characteristics of cases classified as imported cases that were detected on or soon after arrival into Hong Kong from 13 November 2020 through to 31 January 2022, when all arriving persons were required to quarantine in a hotel or a designated quarantine facility. We analysed individual demographics, and clinical information including symptoms and disease severity, virus variants, and Ct values.

**Results:** There were 2269 imported COVID-19 cases aged 0-85 years identified in Hong Kong. Almost half (48.6%) of the imported cases were detected on arrival. A shorter median delay from arrival to isolation was observed in Delta and Omicron cases (3 days) than cases infected with the ancestral strain and other variants (12 days; p<0.001) while lower Ct values at isolation were observed in cases infected with Omicron than the ancestral strain or other variants. No Omicron cases were detected beyond 14 days after arrival, and the cases (n=58, 2.6%) detected after 14 days of quarantine more frequently presented without symptoms at isolation and had a higher RT-PCR Ct-value during isolation. At least some of these cases were post-arrival infections.

**Conclusions:** Testing inbound travellers at arrival and during on-arrival quarantine can detect imported cases early although it may not be sufficient to prevent all introductions of COVID-19 into the community. Public health measures should be adjusted in responses to the emergence of new variants of SARS-CoV-2 based on the epidemiologic evidence from continuous surveillance.

## BACKGROUND

The COVID-19 pandemic and its associated control measures have had an enormous impact on population health and the functioning of societies. Since the emergence SARS-CoV-2 particularly before 2022, many countries have enforced travel restrictions in order to reduce introduction of the virus through inbound travellers.^1^ These measures include arrival and departure restrictions based on residence, vaccination status or other conditions, quarantine for incoming passengers, and COVID-19 test requirements before departure, on arrival, and after arrival. Nevertheless, there has been high variability in application of travel-related measures across jurisdictions.^2, 3^

Hong Kong reported its first COVID-19 case on 23 January 2020 in a returnee from Wuhan.^4^ Since then, travel-related measures have been implemented combined with public health and social measures in the community to control a series of community epidemics.^5^ Measures have undergone changes following the evolution of the pandemic worldwide and the emergence of variants of concerns.^6, 7^ The travel-related measures have substantially reduced the number of virus introductions into the community but at the expense of a 98% reduction in airport passenger traffic and a ban on visitors to the city from most parts of the world between March 2020 and April 2022.

In Hong Kong, hotel quarantines of up to 21 days with frequent testing have been mandatory for international arrivals. Delta and Omicron variants have been found to have a shorter latent period compared to other variants, stimulating discussions about shortening quarantine periods.^8, 9^ In addition, within-quarantine transmission is a risk for individual travellers^10, 11^ that will increase with longer quarantine durations. Understanding the characteristics of imported cases is important for establishing evidence-based control measures. In this study, we characterised imported cases in Hong Kong from the start of mandatory on-arrival hotel quarantine for all arrivals through to 31 January 2022.

## METHODS

Mandatory hotel quarantine was implemented in July 2020 for travellers from high-risk locations^12, 13^ and was extended to all persons arriving in Hong Kong from overseas starting from 13 November 2020. For a short period, arrivals from certain locations were required to quarantine in purpose-built quarantine facilities rather than hotels. We conducted a retrospective cohort study based on a detailed line list without personal data provided by the Hong Kong Department of Health including all confirmed COVID-19 cases and selected for this study imported cases with an arrival from 13 November 2020 through to 31 January 2022. Imported cases were defined as COVID-19 cases that were presumed to have acquired their infection overseas but were confirmed in Hong Kong through reverse transcription polymerase chain reaction (RT-PCR), with an international travel history before the diagnosis. Our study received ethical approval from the Institutional Review Board of the University of Hong Kong.

During the study period, people entering Hong Kong from abroad were tested on arrival and during quarantine by RT-PCR. We divided our study period into five phases based on key changes in testing and quarantine requirements (**Table S1, Table S2**). During our study period, all confirmed cases in Hong Kong were admitted to hospitals or designated facilities for isolation regardless of symptoms or disease severity, including asymptomatic cases and re-positive cases. “Re-positive cases” were defined as individuals who had a previous episode of SARS-CoV-2 infection and tested positive again by RT-PCR, also known as long-term intermittent shedding.^14^ We used information from electronic health records provided by the Hospital Authority to obtain information after case detection, including RT-PCR tests performed during isolation, detection of viral mutations compatible with SARS-CoV-2 variants of concerns, and RT-PCR reaction cycle threshold values (Ct values). Because the date of collection of the first positive RT-PCR before isolation and its Ct-value were not available in the dataset, we instead used the date of isolation and the first Ct-value during isolation as proxies. For cases that tested positive via RT-PCR in quarantine but were negative after transfer to isolation facilities, we imputed a Ct value of 45 at the time of isolation for the cases.^15^

Cases were frequently tested during isolation. Initially, cases would be discharged from isolation if two criteria were met: (i) improvement of clinical conditions and absence of fever in symptomatic cases and (ii) two consecutive negative RT-PCR tests 24 hours apart or tested positive for SARS-CoV-2 IgG.^16^ Discharge criteria were revised in August 2021 requiring symptomatic cases to be isolated for a minimum of 10 days and modifying laboratory criteria: SARS-CoV-2 IgG should be positive and three consecutive PCR tests 24 hours apart with the Ct value of 33 or above.^17^ In October 2021 the criteria were changed again to require two consecutive negative RT-PCR without IgG results needed. Additionally, all cases including asymptomatic and re-positive required a minimum of 10 days of isolation after the first positive test followed by an additional 14 days of isolation in a separate community isolation facility after discharge from the isolation ward.^18^

Imported cases were further characterised into four moments of detection: (i) detected at arrival, (ii) detected during quarantine up to day 14, (iii) detected after day 14 of quarantine, and (iv) detected after arrival with special quarantine arrangements (including imported close contacts of other imported cases that quarantined at designated facilities; aircrew, sea crew and other arrivals with different testing and self-isolation arrangements). We conducted descriptive analyses to summarize case characteristics by moment-of-detection. For individuals detected after day 14 of quarantine, we hypothesised they might be re-positive cases or persons infected after arrival, as it is relatively less likely to observe cases with a very long latent period. Since lower Ct-values during isolation may have increased infectivity and are more compatible with newly incident cases,^19-21^ we further examined the subset of cases detected after day 14 with a minimum Ct-value lower than 30 during isolation. To test for differences across groups we used Chi-squared or fisher tests, and Kruskal-Wallis and Mood’s median test for non-parametric continuous variables. We used Kaplan-Meier curves to compare the time from isolation to a first negative RT-PCR test result based on the presence of symptoms, moment-of-detection, vaccination status and SARS-CoV-2 variant of concern. Cases that did not have a negative test during follow-up were censored at the date of discharge from isolation. We used the log-rank test to test for differences across groups. A p-value <0.05 was used to indicate statistical significance. Statistical analyses were conducted using R version 4.0.3 (R Foundation for Statistical Computing, Vienna, Austria).

## RESULTS

There were 2269 imported COVID-19 cases reported in Hong Kong during the study period, most frequently reported from Southeast Asia and other regions of Asia (**Table S1**) where the Philippines, Indonesia and India were the countries with the highest number of importations (**Figure 1)**. The median age was 35 years ranging from 0 to 85. Over the study period, the highest number of imported cases were observed in the fifth phase of the epidemics (**Table S1, Table S2**) and in January 2022 (**Figure 1**).

**Figure 1.**
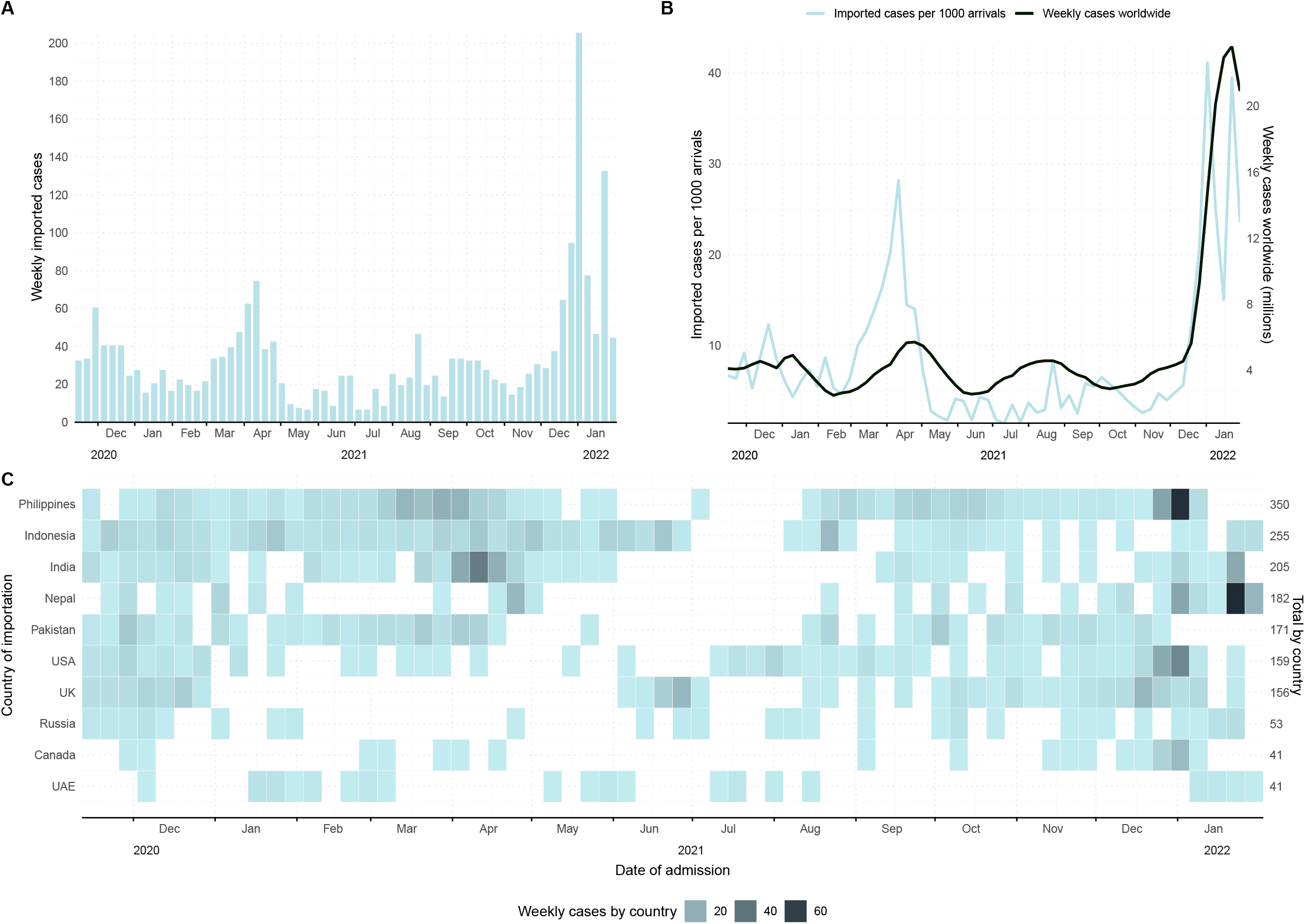
Number of imported COVID-19 cases identified in Hong Kong from 13 November 2020 through 31 January 2022 by week of isolation (A), weekly imported cases per 1000 arrivals to Hong Kong International Airport and weekly cases worldwide (B) and number of imported cases by week of isolation for the most common countries of origin (C).

Half of the imported cases were detected at arrival (48.6%, n=1102), 34.5% (n=782) during quarantine up to day 14 after arrival and 2.6% (n=58) after day 14. For cases not detected on arrival, there was a median delay of 5 days (interquartile range [IQR]: 3-12) between arrival and isolation. The median Ct value at isolation was 29.1 (IQR: 21.2-35.6), higher in asymptomatic cases (30, 22.7-37.9) than in symptomatic cases (23.1, 18.6-29.8, p<0.001).

Among all imported cases, 26.6% were symptomatic at detection, and of these, 19.5% (n=115) reported symptoms before arrival, 23.9% (n=141) on the day of arrival and 56.6% (n=334) after arrival. For cases that initiated symptoms after arrival, the median delay from arrival to onset was 4 days (interquartile range, IQR: 2-6). For cases arriving on or after 12 May 2021 with available information on the vaccination status (n=1122), 650 (57.9%) had received at least two doses of mRNA vaccine, 256 (22.8%) two doses of inactivated vaccine and 216 (19.3%) had other vaccination courses.

Data on SARS-CoV-2 genomic mutations were available for 1295 cases (57.1%), of which 38.4% (497/1295) were compatible with Omicron variants, 38.1% (493/1295) with Delta and 13.1% (170/1295) with other variants of concern (Alpha, Beta or Gamma). Among cases detected after arrival, the median time from arrival to isolation for Omicron and Delta was 3 days (IQR: 3-5) compared to 12 days for other variants (IQR: 5-13; p<0.001) and for the ancestral strain (IQR: 7-13; p<0.001). Among symptomatic cases, the median time from arrival to symptoms onset was shorter for Omicron (3 days, IQR: 2-4) and Delta cases (2 days, 1-5) compared to other variants (7 days, 4-10) and the ancestral strain (5 days, 2-6).

The median Ct-value at isolation for cases infected with Omicron (23, IQR: 19-29) was lower than that for Delta cases (27, 19-34, p<0.001) and cases infected with other variants (29, 22-34; p<0.001) or the ancestral strain (29, 23-34; p<0.001) (**Figure 2**). This value changed with the delay from arrival to isolation and was consistently lower for Omicron cases (**Figure 3)**. The median minimum Ct-values during isolation also differed across variants, with Omicron cases (20, 18-24) lower than Delta (22, 18-30; p<0.001), other variants (27, 20-31; p<0.001) and the ancestral strain (25, 20-31; p<0.001) (**Figure 2**).

**Figure 2.**
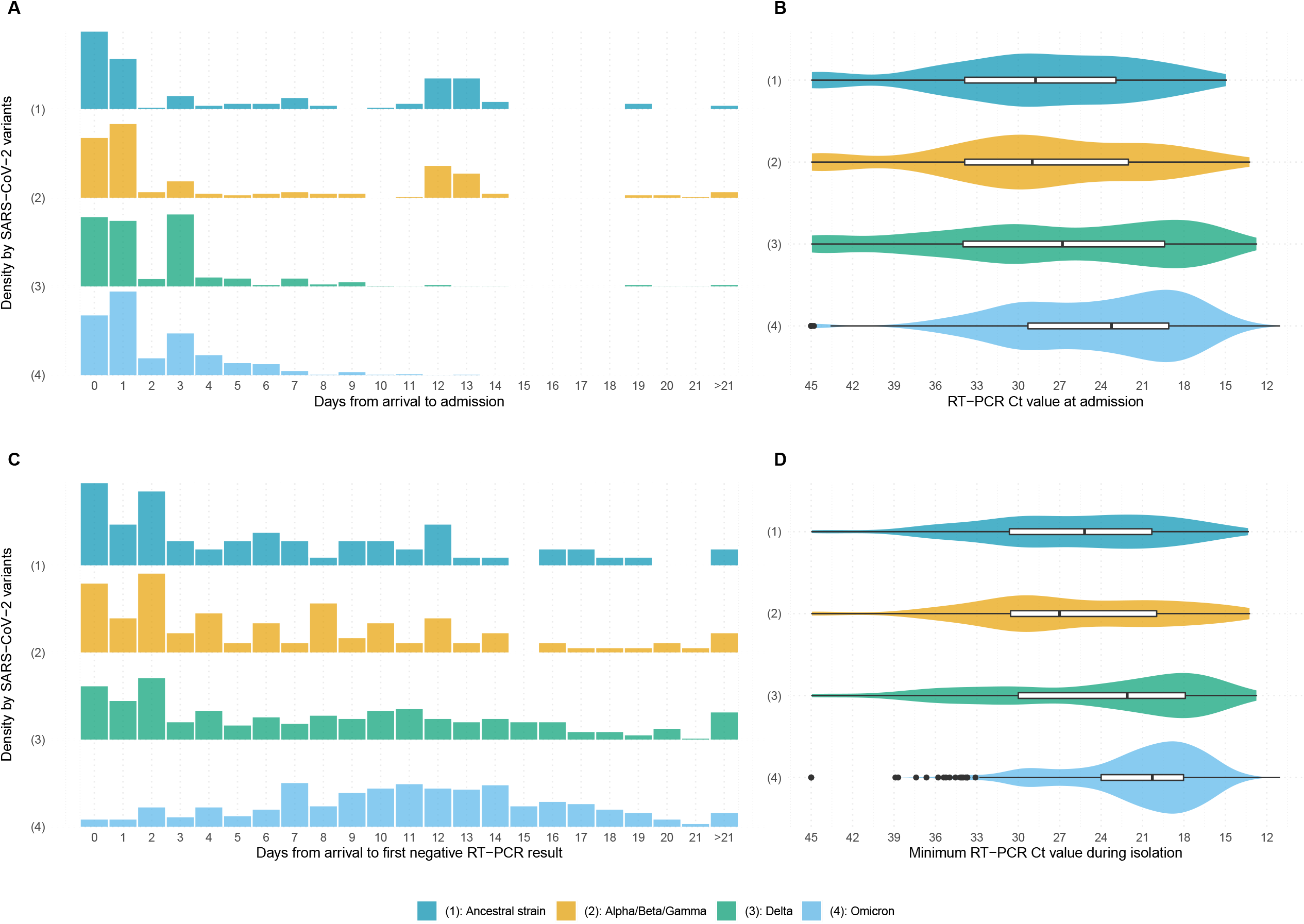
Differences across SARS-CoV-2 variants among imported COVID-19 cases identified in Hong Kong in days from arrival to admission for isolation (A), RT-PCR Ct-value at admission for isolation (B), days from isolation to negative RT-PCR (C) and minimum RT-PCR Ct-value during isolation (D).

**Figure 3.**
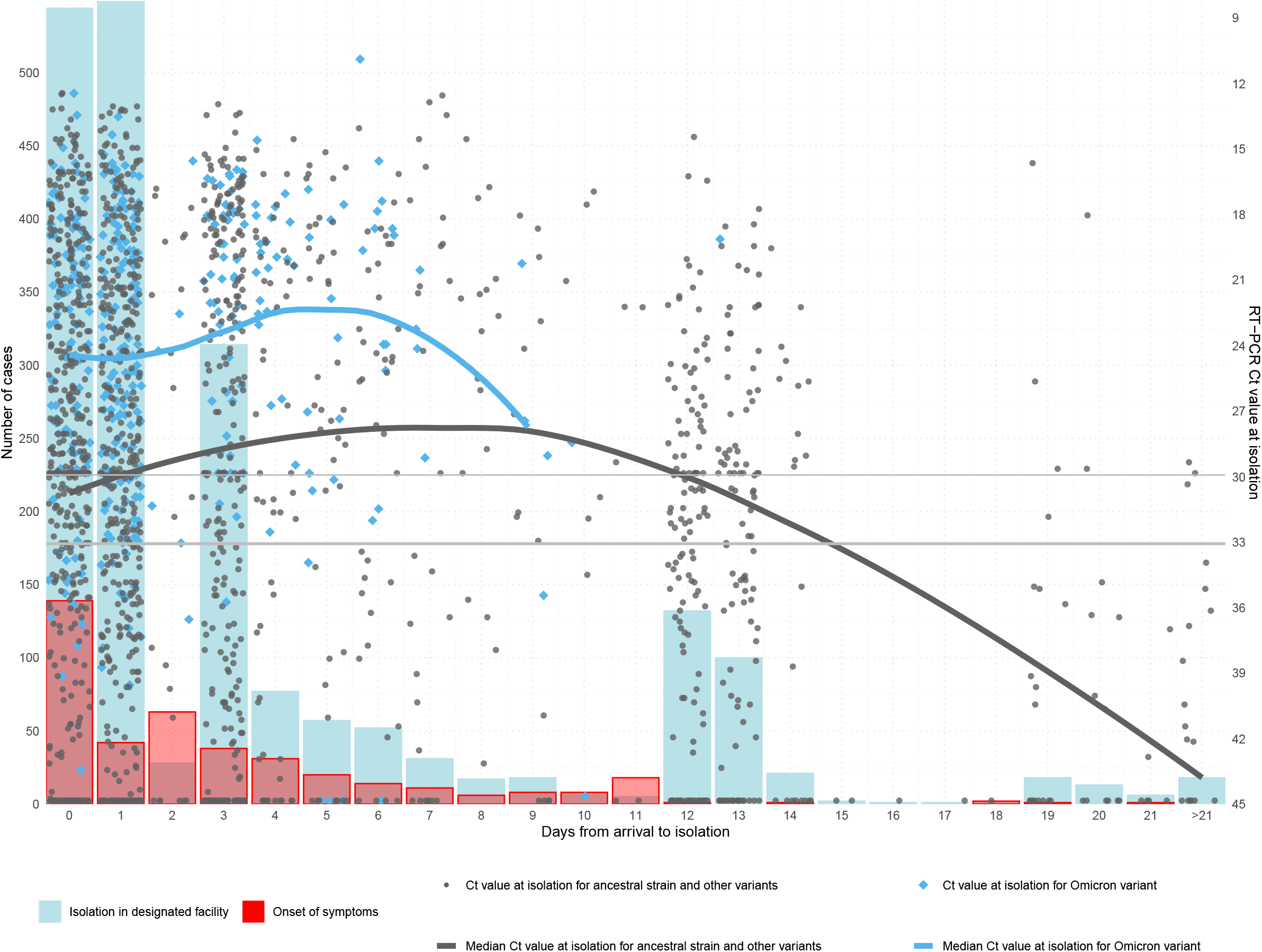
Number of imported cases with Omicron and other SARS-CoV-2 variants identified in Hong Kong by day from arrival to isolation and day from arrival to symptom onset, and RT-PCR Ct-value at isolation of the imported cases by day from arrival to admission for isolation. Horizontal lines indicate RT-PCR Ct-value at 33 (samples with very low viral load, the criterion used for release of isolation during specific time periods) and 30 (samples with low viral load, indicating a possible re-positive result i.e. long-term intermittent shedding).

Fifty-eight (2.6%) cases were detected after day 14. These cases were more frequently female, mostly detected during the second phase (21-day hotel quarantine for all international arrivals, **Table S2**) and the most common region of importation was Southeast Asia (60.3%). Only 10.3% of them were symptomatic compared to 32.0% detected up to day 14 and 22.8% detected at arrival (p<0.001). Their median Ct value at isolation (44, IQR: 36-45) was higher than in the cases detected within 14 days (30, 22-38, p<0.001) and those detected at arrival (28, 21-34, p<0.001). The median minimum Ct value during isolation was 24 (19-30), 27 (20-35) and 36 (31-45) for cases detected at arrival, within and after 14 days of quarantine, respectively (p<0.001). No Omicron case was detected after day 14 (**Table 1**).

**Table 1.** Characteristics of imported cases in Hong Kong by moment of detection (excluding cases with special quarantine arrangements)

|  | At arrival<br>(N=1102) | Up to day 14 of<br>quarantine<br>(N=782) | After day 14 of<br>quarantine<br>(N=58) | P-value |
| --- | --- | --- | --- | --- |
| Age group |  |  |  |  |
| <20 years | 103 (9.35%) | 84 (10.7%) | 7 (12.1%) | 0.470 |
| 20-39 years | 577 (52.4%) | 421 (53.8%) | 31 (53.4%) |  |
| 40-59 years | 350 (31.8%) | 221 (28.3%) | 14 (24.1%) |  |
| >59 years | 72 (6.53%) | 56 (7.16%) | 6 (10.3%) |  |
| Sex |  |  |  |  |
| Female | 574 (52.1%) | 460 (58.8%) | 44 (75.9%) | <0.001 |
| Male | 528 (47.9%) | 322 (41.2%) | 14 (24.1%) |  |
| Region of importation |  |  |  |  |
| Asia (not including Southeast Asia) | 330 (30.3%) | 282 (36.4%) | 15 (25.9%) | <0.001 |
| Southeast Asia | 280 (25.7%) | 280 (36.2%) | 35 (60.3%) |  |
| Europe | 255 (23.4%) | 117 (15.1%) | 4 (6.90%) |  |
| Others | 223 (20.5%) | 95 (12.3%) | 4 (6.90%) |  |
| Period of arrival (% by row) |  |  |  |  |
| First phase (N=258) | 187 (72.5%) | 68 (26.4%) | 3 (1.2%) | <0.001 |
| Second phase (N=561) | 282 (50.3%) | 239 (42.6%) | 40 (7.1%) |  |
| Third phase (N=185) | 100 (54.1%) | 83 (44.9%) | 2 (1.1%) |  |
| Fourth phase (N=324) | 158 (48.8%) | 154 (47.5%) | 12 (3.7%) |  |
| Fifth phase (N=614) | 375 (61.1%) | 238 (38.8%) | 1 (0.2%) |  |
| Vaccination status (from 12 May 2021) |  |  |  |  |
| 2 doses mRNA | 342 (54.0%) | 202 (42.5%) | 2 (13.3%) | <0.001 |
| 2 doses inactivated | 93 (14.7%) | 124 (26.1%) | 6 (40.0%) |  |
| Other vaccination courses | 115 (18.2%) | 72 (15.2%) | 4 (26.7%) |  |
| Not vaccinated/Not reported | 83 (13.1%) | 77 (16.2%) | 3 (20.0%) |  |
| Presence of symptoms |  |  |  |  |
| Asymptomatic | 851 (77.2%) | 532 (68.0%) | 52 (89.7%) | <0.001 |
| Symptomatic | 251 (22.8%) | 250 (32.0%) | 6 (10.3%) |  |
| Variants of concern |  |  |  |  |
| Omicron | 273 (24.8%) | 117 (15.0%) | 0 (0%) | <0.001 |
| Delta | 245 (22.2%) | 164 (21.0%) | 9 (15.5%) |  |
| Other variants (Alpha/Beta/Gamma) | 85 (7.71%) | 59 (7.54%) | 7 (12.1%) |  |
| Ancestral strain | 65 (5.90%) | 55 (7.03%) | 4 (6.90%) |  |
| Not tested/Indeterminate | 434 (39.4%) | 387 (49.5%) | 38 (65.5%) |  |
| RT-PCR Ct-value at isolation |  |  |  |  |
| Mean (SD) | 28.2 (8.73) | 30.2 (9.79) | 39.8 (6.98) | <0.001 |
| Median [Q1, Q3] | 27.7 [21.1, 33.8] | 30.0 [21.5, 38.5] | 44.0 [36.1, 45.0] |  |
| Minimum RT-PCR Ct-value during isolation |  |  |  |  |
| Mean (SD) | 25.4 (8.22) | 27.9 (9.04) | 36.3 (7.43) | <0.001 |
| Median [Q1, Q3] | 23.6 [18.7, 30.5] | 27.3 [19.9, 34.8] | 36.1 [31.3, 44.6] |  |
| RT-PCR Ct-value ever lower than 30 during isolation |  |  |  |  |
| No | 334 (30.4%) | 336 (43.2%) | 49 (84.5%) | <0.001 |
| Yes | 766 (69.6%) | 441 (56.8%) | 9 (15.5%) |  |

After excluding cases without available data or under special quarantine arrangements, among 8 cases detected beyond 14 days of quarantine with at least one Ct-value lower than 30 during isolation, three were probably infected during hotel quarantine,^11^ three were suspected cases of within-hotel transmission (epidemiological overlap but genomic sequences not available or inconclusive),^11^ one was related to an air travel-related outbreak but within-quarantine transmission cannot be ruled out,^22^ and another was classified as a re-positive case^23^ (**Table S3)**.

Among the 5 symptomatic cases with data available detected after day 14, three had a Ct-value lower than 30 during isolation (**Table S3**). The other two correspond to a young child who had symptom onset on day 9 after arrival but was only isolated on day 22, and to a case arriving from the Philippines who had symptom onset 21 days after arrival, with a low viral load when first detected and then an indeterminate PCR result at the isolation facility, consistent with detection of a re-positive case and co-incidental symptoms from a separate infection.

Patients were isolated for a median of 12 days (IQR: 8-17). On 23 March 2022, 40 cases (1.8%) were still in isolation, and two died. Among currently isolated or already discharged cases, only 6 had severe or serious disease. The median time from isolation to the first PCR negative result was 10 days (95% confidence interval, CI: 10, 11) compared to 5 days (95% CI: 4,5) from isolation to the first Ct value at 33 or above. The time from isolation to the first PCR negative result was shorter for asymptomatic patients (median: 9 days, 95% CI: 9, 10) compared to symptomatic ones (median: 12 days, 95% CI: 12, 13) and for individuals detected after 14 days of quarantine (median: 0.5 days, 95% CI: 0, 2), compared to cases detected at arrival (median: 12 days, 95% CI: 11-13), and up to day 14 of quarantine (median: 9 days, 95% CI: 8, 9). The time from isolation to the first PCR result with Ct value at 33 or above was similar among cases with SARS-CoV-2 variants and the ancestral strain (**Figure 4**).

**Figure 4.**
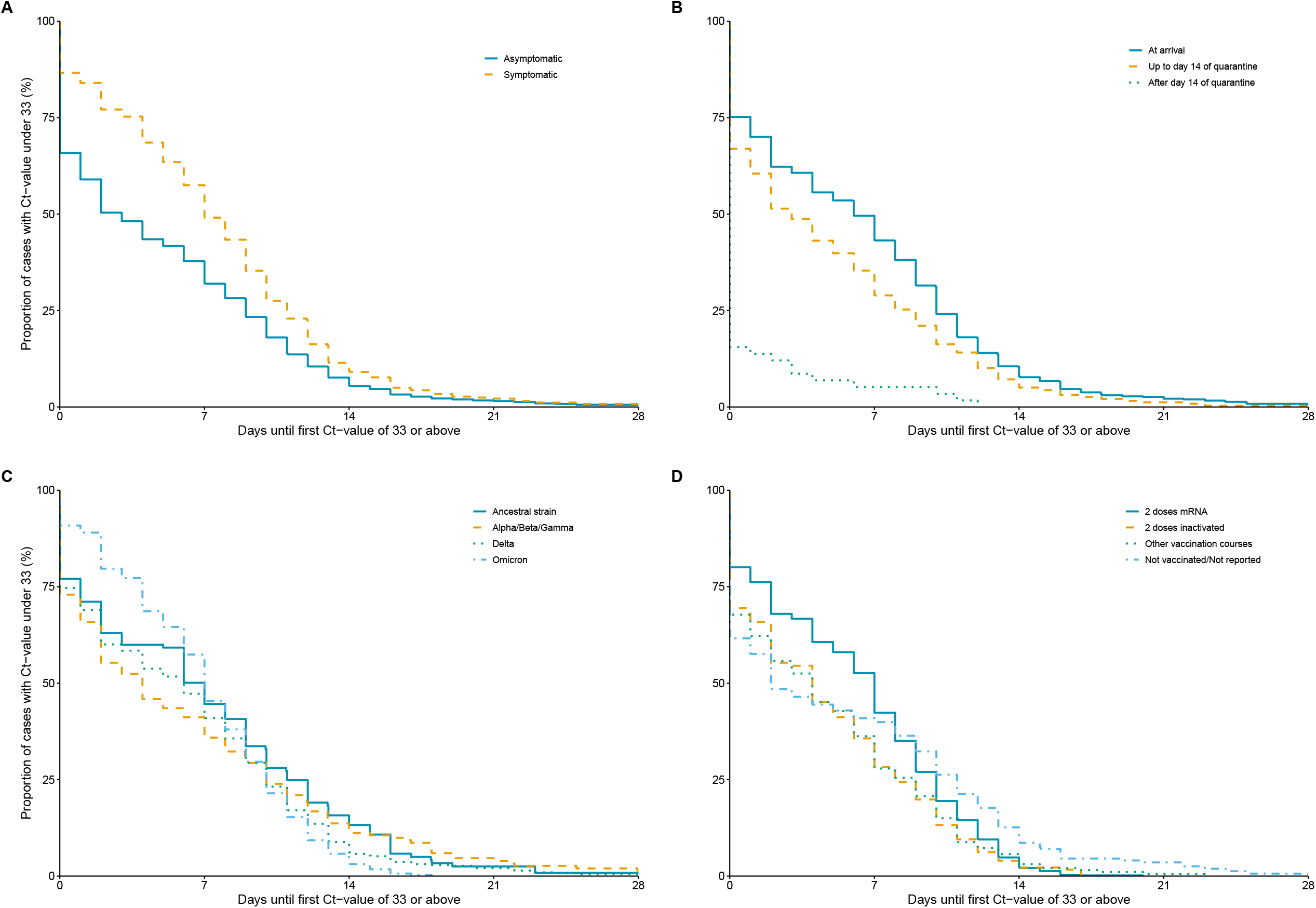
Kaplan-Meier curves for the time from isolation to first RT-PCR result with Ct value at 33 or above among imported COVID-19 cases identified in Hong Kong by: (A) presence of symptoms; (B) moment of detection; (C) type of SARS-CoV-2; (D) vaccination status including receipt of mRNA vaccination (BNT162b2), inactivated vaccine (CoronaVac), or other vaccine types.

## DISCUSSION

Our analysis on 2269 imported cases in Hong Kong over 18 months indicated strict on-arrival measures could reduce community introductions of the virus. The imported cases were largely asymptomatic at confirmation and presented mild symptoms during hospital isolation.

The majority of imported infections could be detected at arrival or within a few days after arrival, with some cases detected beyond 14-day quarantine likely due to re-positivity or transmission during quarantine. Differences in time from arrival to onset and detection across SARS-CoV-2 variants were apparent.

The relatively shorter median delay from arrival to symptom onset in imported Omicron and Delta cases than cases of the ancestral strains and other variants is consistent with the findings from a systematic review showing a mean incubation period of 6.3 days for infections of the ancestral strain compared to 4.8 and 3.6 days for Delta and Omicron, respectively.^24^ Studies from China estimated a mean latent period of 5.5 days for SARS-CoV-2 in local outbreaks before April 2021^25^ and a shorter latent period for Delta (mean: 3.9 days).^26^ Our estimates of the median time from arrival to isolation in imported cases also suggested a difference between cases infected with Delta and Omicron (median: 3, third quartile: 5 days) and with other variants and ancestral strains (median: 12 days, third quartile: 13 days). Given the circulation of SARS-CoV-2 in origin countries,^1, 27^ the duration of quarantine for inbound travellers should be adjusted in response to the changes in epidemiological characteristics of the virus.

In our study, some cases mostly asymptomatic were detected after 14 days of quarantine, though they often had higher Ct values (median Ct-value at isolation: 44.0, minimum Ct value during isolation: 36.1) compared to those detected earlier, indicating a lower viral burden and perhaps less transmissibility^21^. It has been widely documented that positive RT-PCR tests might occur weeks after recovery for SARS-CoV-2 sometimes following previous negative results.^19, 28-30^ This is compatible with the hypothesis that re-positive cases who recovered from previous infections before departure would be very unlikely to be contagious.^31^ Conversely, some cases with a negative RT-PCR result at arrival were detected during or after quarantine with lower Ct values were more likely due to infections acquired during quarantine (e.g. among families traveling together, or through transmission between rooms in a quarantine hotel) rather than long incubation periods.^11^

Transmission events occurring between rooms in quarantine hotels have been reported in Hong Kong, Australia, and New Zealand, among others, and have resulted in COVID-19 reintroductions in jurisdictions that pursued an elimination strategy.^10, 11, 32-34^ Hotel rooms are not designed for quarantine purposes and may have limitations in the ventilation systems or floor plan design.^33, 35^ Hong Kong experienced a large epidemic of Omicron BA.2 in early 2022 which had originated in a quarantine hotel transmission between arriving persons staying in different hotel rooms.^36-38^

This study has several limitations. First, we did not have the date of sample collection of the diagnostic RT-PCR to determine the moment of detection, instead, we used the date of isolation in designated facilities. It is possible that for some cases the moment of detection occurred a few days before isolation, but we believe these were a minority and would therefore have limited impact on estimates. Second, different RT-PCR testing, quarantine, and discharge criteria across periods together with changes in arrival characteristics may confound the relationship between key parameters and SARS-CoV-2 variants. Although in our study, clear differences across variants that align with scientific literature were described, caution is still needed in their interpretation.

In conclusion, testing inbound travellers at arrival and during on-arrival quarantine could detect imported cases early although it might not be sufficient to prevent all introductions of SARS-CoV-2 into the community. Travel-related public health measures should be implemented with a holistic consideration of the potential individual and societal costs as well as the potential public health benefits through reducing the rate of community importations of infections. Travel-related measures may need to be adjusted in response to the epidemiologic parameters of new variants of SARS-CoV-2.

## Data Availability

All data produced in the present study are available upon reasonable request to the authors.

## ACKNOWLEDGMENTS

The authors acknowledge the Hospital Authority of Hong Kong and the Centre for Health Protection of the Hong Kong Department of Health for providing epidemiological data for the study. The authors thank Julie Au for technical support.

## DATA AVAILABILITY

The data analysed in this study were provided by the Hospital Authority of Hong Kong and the Centre for Health Protection of the Hong Kong Department of Health, and cannot be shared without the permission from the two parties.

## AUTHOR CONTRIBUTIONS

MM-S, PW and BJC conceived the study. MM-S, DCA, YL, EHYL collected the data. MM-S conducted the analysis and wrote the first draft of the manuscript. All authors critically reviewed, revised, and commented on the manuscript and approved the final version of the manuscript.

## SUPPLEMENTARY MATERIAL

**Table S1.** Characteristics of imported cases that arrived in Hong Kong from 13 November 2020 to 31 January 2022.

|  | **Overall**<br>**(N=2269)** |
| --- | --- |
| **Age group** |  |
| <20 years | 251 (11.1%) |
| 20-39 years | 1182 (52.1%) |
| 40-59 years | 678 (29.9%) |
| >59 years | 157 (6.92%) |
| **Sex** |  |
| Female | 1218 (53.7%) |
| Male | 1051 (46.3%) |
| **Region of importation** |  |
| Asia (not including Southeast Asia) | 781 (34.9%) |
| Southeast Asia | 652 (29.1%) |
| Europe | 426 (19.0%) |
| North America | 207 (9.24%) |
| Africa | 59 (2.63%) |
| South America | 13 (0.580%) |
| Multiple regions | 103 (4.60%) |
| **Period of arrival\*** |  |
| First phase<sup>a</sup> | 277 (12.2%) |
| Second phase<sup>b</sup> | 608 (26.8%) |
| Third phase<sup>c</sup> | 221 (9.74%) |
| Fourth phase<sup>d</sup> | 385 (17.0%) |
| Fifth phase<sup>e</sup> | 778 (34.3%) |
| **Presence of symptoms** |  |
| Asymptomatic | 1664 (73.4%) |
| Symptomatic | 603 (26.6%) |
| **Moment of symptom onset** |  |
| Before arrival | 115 (19.5%) |
| Day of arrival | 141 (23.9%) |
| After arrival | 334 (56.6%) |
| **Vaccination status (from 12 May 2021)** |  |
| 2 doses mRNA | 650 (47.0%) |
| 2 doses inactivated | 256 (18.5%) |
| Other vaccination courses | 216 (15.6%) |
| Not vaccinated/Not reported | 262 (18.9%) |
| **Moment of detection** |  |
| At arrival | 1102 (48.6%) |
| Up to day 14 of quarantine | 782 (34.5%) |
| After day 14 of quarantine | 58 (2.56%) |
| After arrival, special quarantine arrangements | 326 (14.4%) |

<sup>a</sup>14-day hotel quarantine (13 November-24 December 2020), <sup>b</sup>21-day hotel quarantine (25 December 2021-11 May 2021), <sup>c</sup>7,14 or 21-day hotel quarantine based on vaccination status and country of arrival (12 May-19 August 2021), <sup>d</sup>14 or 21-day hotel quarantine based on vaccination status and country of arrival (20 August-27 November 2021), <sup>e</sup>14 or 21-day hotel quarantine based on vaccination status and country of arrival, enhanced surveillance of arrivals from high risk countries (28 November 2021-31 January 2022)
\*Details in Table S1

**Table S2.**
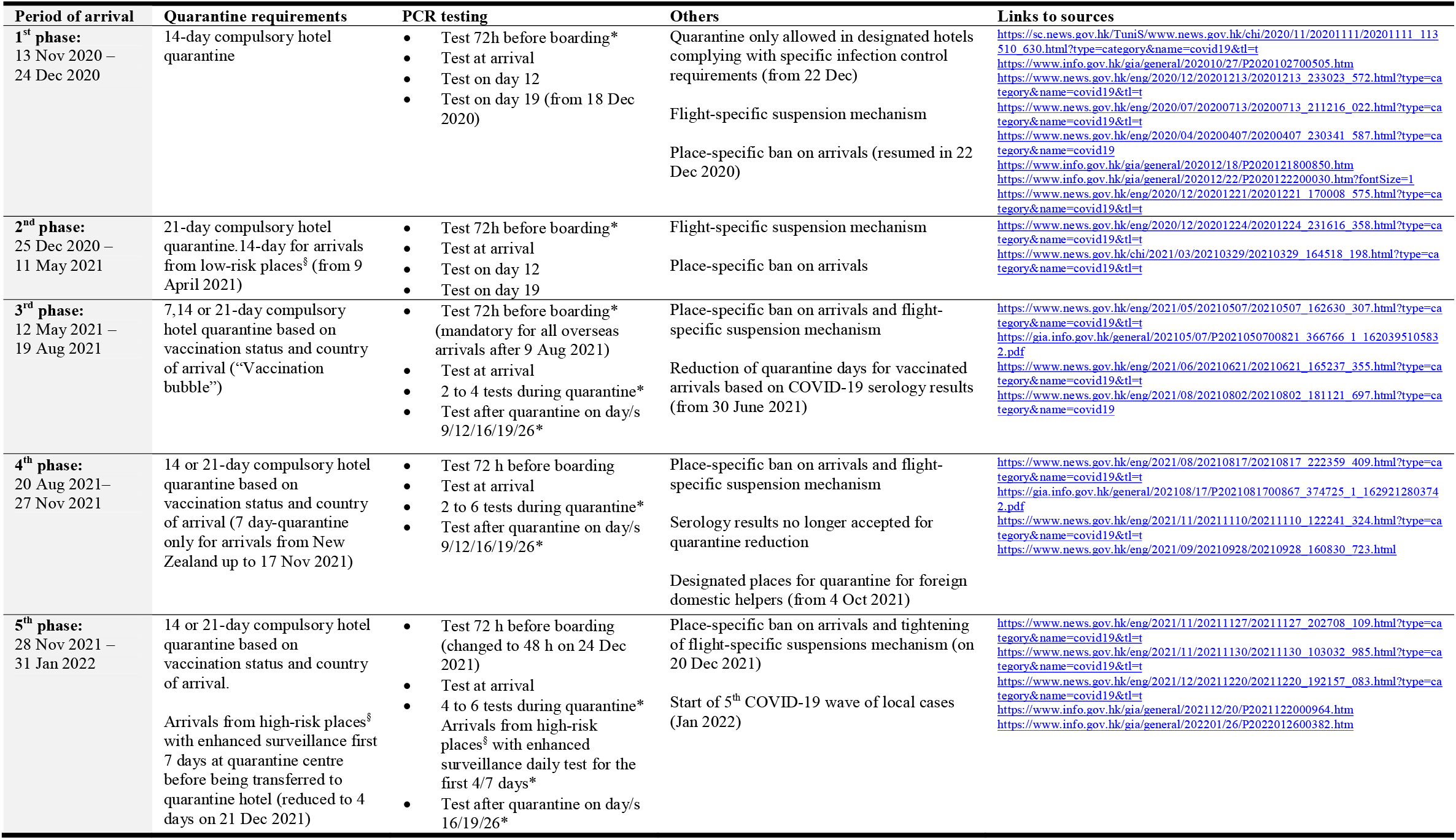
Testing and quarantine requirements for international arrivals in Hong Kong, 13 November 2020 to 31 January 2022

**Table S3.** Description of cases detected after 14 days of quarantine with Ct-values lower than 30 during isolation

| Age group, years | Month of arrival | Country of origin | Days from arrival to isolation | Mandated quarantine length | Vaccination status | Days from arrival to onset | Lowest RT-PCR Ct-value during isolation | Case details<sup>11, 22, 23</sup> |
| --- | --- | --- | --- | --- | --- | --- | --- | --- |
| 40-59 | Dec 2020 | Nepal | 20 days | 21 days | NA | 18 days | 18.2 | Suspected transmission during hotel quarantine<sup>a</sup> |
| >59 | Feb 2021 | Canada | 19 days | 21 days | NA | 18 days | 15.8 | Probable transmission during hotel quarantine<sup>b</sup> |
| 30-39 | March 2021 | Indonesia | 20 days | 21 days | NA | 19 days | 29.4 | Probable transmission during hotel quarantine<sup>b</sup> |
| 30-39 | April 2021 | India | 21 days | 21 days | NA | Asymptomatic | 26.8 | Part of an air-travel-related cluster of 59 cases arriving on a single flight from New Delhi. Transmission during quarantine cannot be ruled out |
| 40-49 | March 2021 | Philippines | 33 days | 21 days | NA | Asymptomatic | 26.9 | Suspected transmission during hotel quarantine<sup>a</sup>. Detected after hotel quarantine through required community testing for foreign domestic helpers |
| 40-49 | June 2021 | Indonesia | 19 days | 21 days | Not vaccinated/not reported | Asymptomatic | 25.8 | Probable transmission during hotel quarantine<sup>b</sup> |
| <20 | October 2021 | Italy, Qatar | 26 days | 21 days | Not vaccinated/not reported | Asymptomatic | 29.5 | Case classified as re-positive |
| >59 | October 2021 | Thailand | 19 days | 21 days | 2 doses inactivated vaccine | Asymptomatic | 19.6 | Suspected transmission during hotel quarantine<sup>a</sup>. |

NA: not applicable, <sup>a</sup>Epidemiological overlap with an imported case staying in a different room of the same quarantine hotel but genome sequences not available or inconclusive <sup>b</sup>Epidemiological overlap and identic genomic sequencing or $\leq 4$ genome-wide substitutions, in cases with different countries of departure

